# Relation of Doubling Time and Reproduction Number with Testing Rate for Corona Infections in India

**DOI:** 10.1101/2020.08.29.20184283

**Authors:** Abhaya Indrayan, Aman Mishra

**Affiliations:** Department of Clinical Research, Max Healthcare, Delhi

## Abstract

There is a general feeling that increased testing has a salutary effect on the course of the current coronavirus epidemic. We quantify the relationship of testing rate with the doubling time and reproduction number in India, estimate the effect, and use these relationships to make projections for the near future.

## Introduction

First 1000 cases of corona infections in India accumulated on April 3, 2020. The caseload since then has steeply increased to more than 3 million as of August 23, 2020. Testing was low in the beginning with just about 0.06 per 1000 population but has made rapid strides to nearly 25 per 1000 population on August 23, 2020. There is a general belief that increased testing helps in controlling the epidemic. Considerable data are now available to study the trend of cases, doubling time, reproduction number, and testing rate, to examine how far they are related, and what lessons, if any, can be drawn with the experience we have of this epidemic in India thus far, with particular reference to the role of increased testing.

## Material and Methods

The cumulative total of the daily cases and tests were downloaded from the crowd sourced OurWorldinData^1^. Our random checks for specific dates reveal that the numbers in this database match the official data from time to time at the Ministry of Health and Welfare site. Doubling time was calculated from the data starting from April 3, 2020, when the number of cases was at least 1000. Before that the trend was erratic. An ingenious method, as mentioned in a short while, was used to estimate the reproduction number from the doubling time.

Reproduction number is the number of secondary infections generated by an infected person over the entire infectious period. This defines transmissibility and epidemic potential. This is calculated at baseline at the beginning of an epidemic and at different stages subsequently. The basic reproduction number (*R*_0_) is the number of people infected by one infected person over the course of the infectious period when s/he is in a community where everybody is susceptible. This is used in the initial phase of the epidemic when almost all are susceptible and cannot be altered. As the epidemic progresses and the infection spreads, immunity develops in some people and the pool of susceptibles declines. Also, steps are taken to control the epidemic. Thus, in the subsequent stages, reproduction number (*R_t_*) is calculated at the time *t*, depending on the situation at that time, and has varying names such as effective reproduction number and time-dependent reproduction number. This can be used to evaluate effectiveness of the current public health interventions.

An intricate method based on likelihood^2^ is generally suggested for calculating the reproduction number. This requires assumptions regarding parameters such as infectious period, serial interval, and generation interval. The estimates vary from study to study due to varying assumptions. However, we used a simple method based on the doubling period and exponential growth and calling it a period reproduction number. Both the parameters used in this calculation do not have any subjective element and will not vary from study to study.

An essential requirement for direct calculation of the reproduction number is the infectious period. According to Housen et al.^3^, the infectious period of corona may start 1 to 3 days before the onset of symptoms and may continue for 7 days after the symptoms begin. The Centre of Disease Control^4^ found that persons with mild to moderate disease remain infectious no longer than 10 days after the onset of symptoms. Byrne et al.^5^ reported post-symptomatic infectious period ranging from 2 to 22 days. In view of such varying reports, we decided to work with an assumed infectious period of 10 days to calculate the period reproduction number. Ten days looks like the median infectious period – median being the appropriate representative in view of right skewed distribution of the infectious period (Byrne) as are almost all durations.

If the doubling time is *D* days, daily compounding gives

the number of new infected persons in *I* days per infected person = 2*^I^*^/^*^D^*.

We are calling it the period reproductive number. This is not the same as generally cited in the literature based on intricate calculation^6^ with R package but has nearly the same meaning. The period reproduction number can be easily estimated from the doubling time at different stages of the epidemic.

The relationship of doubling time and reproduction number with testing rate was studied by trying several forms of regressions. The one that looked close to the observed data and gave a high value of *R*^2^ was considered right for further exploration.

## Results

The progress of the epidemic in India with doubling dates (Figure 1 and Table 1) shows how the doubling time has increased over time. It was only 3 days in the beginning on April 6, 2020, but steadily increased to nearly 21 days in the second week of August 2020. This has remained at this level since then by the end of August 2020 when this communication was prepared.

**Figure 1.**
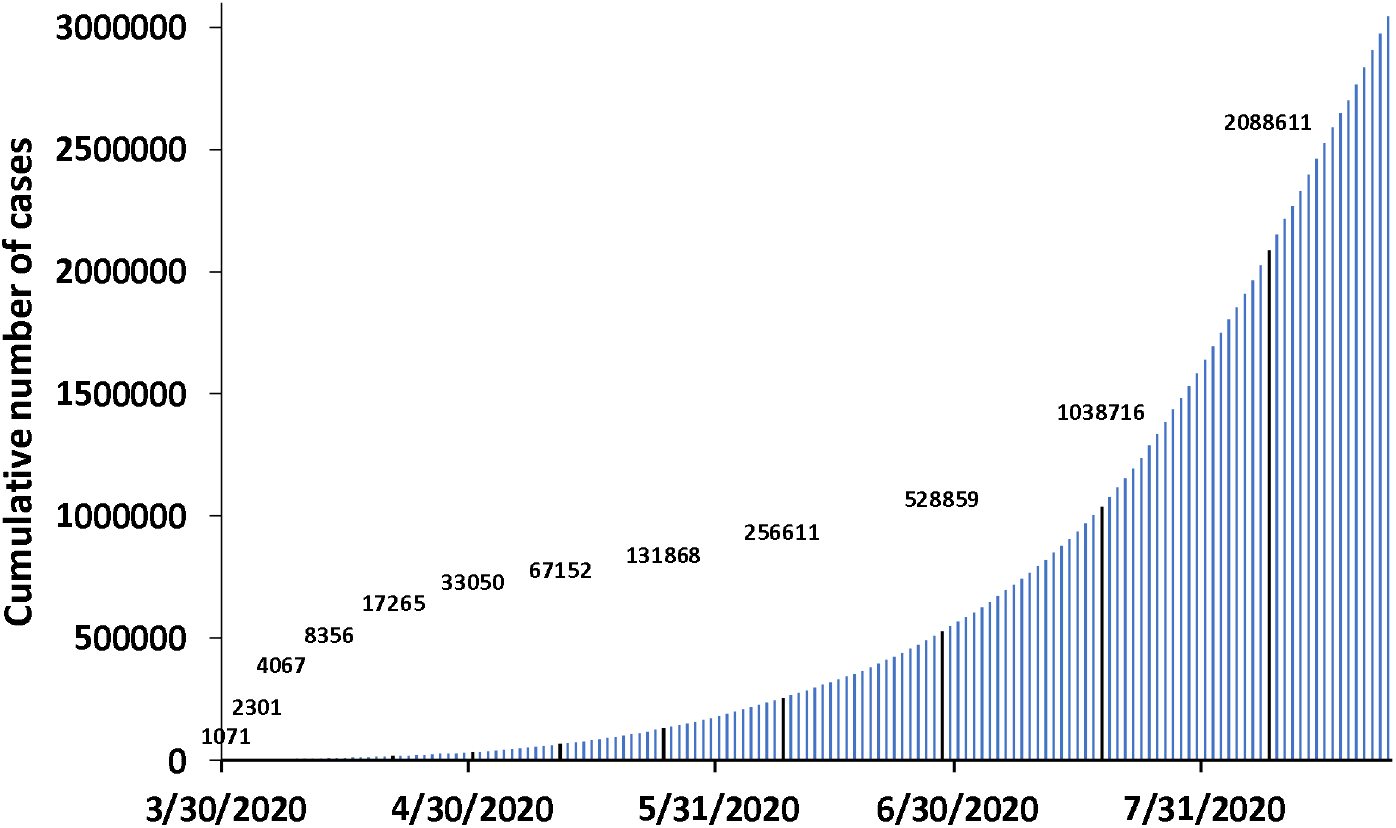
Doubling time of the cumulative cases

**Table 1.** Doubling time, period reproduction number and cumulative tests per 1000 population

| Date | Doubling time (days) | Period reproduction number ($R_0$) | Cumulative tests per thousand population |
| --- | --- | --- | --- |
| 3 Apr 2020 |  | 5.66 | 0.058 |
| 6 Apr 2020 | 3 | 10.08 | 0.083 |
| 12 Apr 2020 | 6 | 3.17 | 0.158 |
| 20 Apr 2020 | 8 | 2.38 | 0.335 |
| 30 Apr 2020 | 10 | 2.00 | 0.654 |
| 11 May 2020 | 11 | 1.88 | 1.275 |
| 24 May 2020 | 13 | 1.70 | 2.198 |
| 8 Jun 2020 | 15 | 1.59 | 3.562 |
| 28 Jun 2020 | 20 | 1.41 | 6.086 |
| 18 Jul 2020 | 20 | 1.41 | 9.994 |
| 8 Aug 2020 | 21 | 1.39 | 17.468 |

The derived data for the dates with the doubling time, period reproduction number and the tests per 1000 as of August 23, 2020, are in Table 1. Further doubling did not occur till the time of this study. The Pearson’s correlation coefficient of doubling time with reproduction number was −0.73, between doubling time and tests per 1000 was +0.83, and between reproduction number and tests per 1000 was −0.42. These correlations measure only the linear relationship. The trends of these numbers over time (Figure2) clearly show that the relationships were not linear – thus Pearson’s correlation is not an adequate measure of relationship in this case.

**Figure 2.**
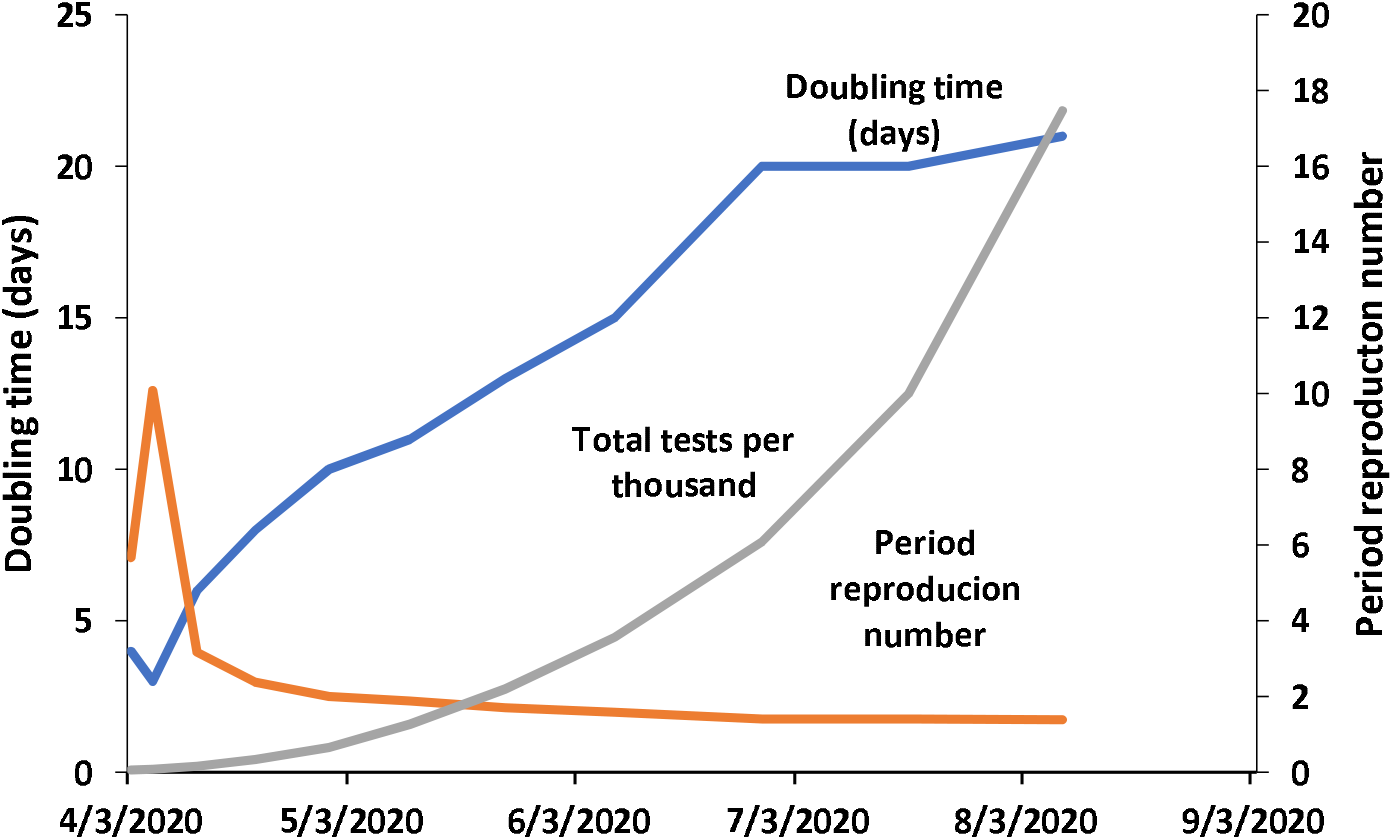
Trend of doubling time, tests, and period reproductive number

We explored several regressions to express the relationship of doubling time and reproduction number with tests per 1000 population. Among these, the best fit for doubling time was the following logarithmic equation:

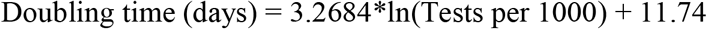

This gave *R^2^=* 0.97, which is high by any standard and the fit is extremely good (Figure 3). Such a high R^2^ tells us that the doubling time was intimately related to logarithmic of the number of tests per 1000 population. The effect of increased testing on doubling time depended on the existing level of testing. However, this is just a relationship and does not express causality. Other factors such as lockdown, face masks, and social distancing may have a more significant role which are not included in our exercise.

**Figure 3.**
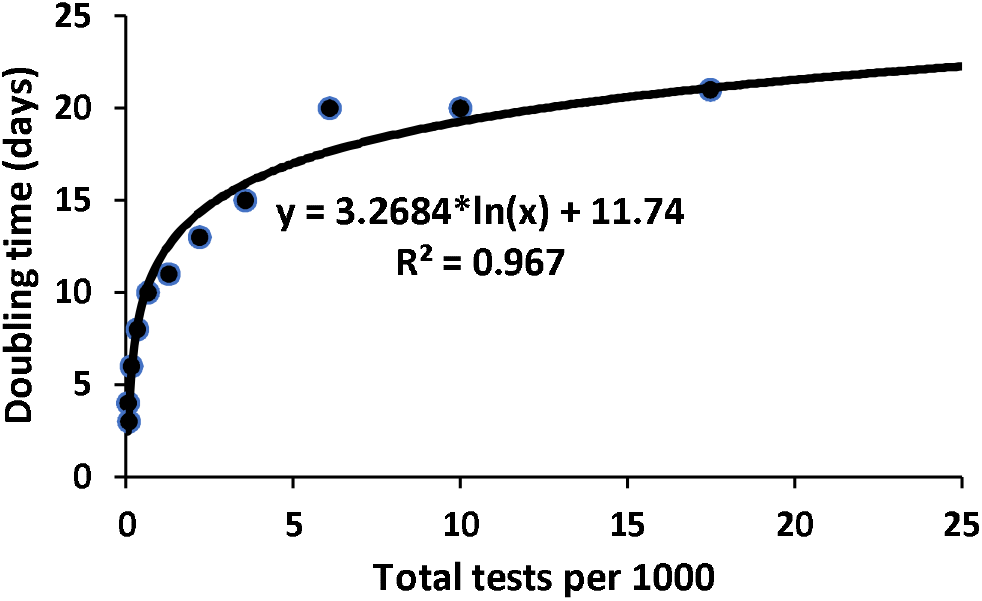
Regression fit of doubling time on tests per 1000 population

The second relationship we examined is between period reproduction number and the tests. Among those we tried, the best regression for this was

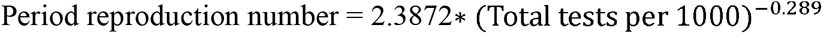

With an *R^2^=* 0.78, this is not as close fit as for doubling time but is still reasonably good (Figure 4). This also is far from linear. This equation says that the period reproduction number was related to the negative power of the number of tests per 1000 population. If the tests rise to 25 per 1000 population, the doubling time is likely to be around 22.3 days and the period reproduction number nearly 0.94. If the present trend continues, the reproduction number can become 1.0 when testing is 20.3 per 1000 population. This reproduction number corresponds to a stable epidemic but not the end of the epidemic.

**Figure 4.**
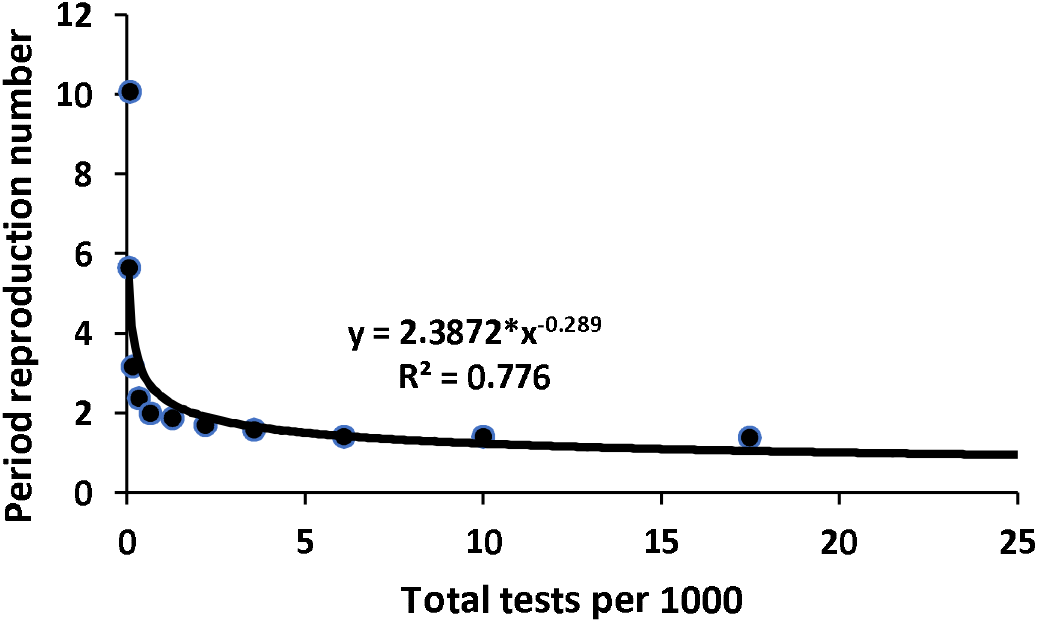
Regression fit for period reproduction number on tests per 1000 population

## Discussion

No comparable study was available on relationship of doubling time and reproduction number with the testing rate. An extremely good equation relating doubling time with testing rate and a reasonably good equation on relation of reproductive number and testing rate have been obtained in our study. The results provide evidence that the doubling time and reproduction number are intimately related with the testing rate in India. This is not a cause-effect relationship since well-known measures of lock down, wearing face masks, social distancing, and avoiding gathering were also operative at the same time and may have handsomely contributed. Yet, the equations we obtained provide an idea of how the testing rates may have affected the course of the epidemic and document the kind of relationship doubling time and reproduction rate may have with the testing rates.

For experts, this study has several shortcomings. For quality research, these should be clearly stated in a transparent manner^7^. The study assumes an infectious period of 10 days which of course varies from person to person and location to location. This period has a right skewed distribution^5^ and has been reported differently by different workers. Perhaps no definite information is available for the infectious period of coronavirus. We are taking a simplistic view and using 10 days, which could be the median infectious period. However, such an assumption may not be much of an impediment for examining the effect testing rates on the doubling time and the reproduction number because their pattern will remain the same even if the infectious period is different.

Second, the number of cases is based on the reporting that we all know is deficient everywhere in the world, including Sweden^8^. The undetected cases are not known and no correction factor is available. Almost all researchers have worked with the available data and we have also done so. Many previous publications have not even considered this as a limitation.

Third, testing includes the RT-PCR test, which is considered the gold standard, but also, particularly recently, the rapid antigen tests which are known to have substantial false negative results^9^. However, our focus in this communication is on testing per se without considering the type of test.

Fourthly, we are using doubling time to calculate the period reproduction number. Our calculations are for the rate of increase of infections in the infectious period per infected person. This is the improvisation we are suggesting as a replacement of the intricate calculations of reproduction number. Our method found a reproduction number of 5.66 in the first week of April compare to 1.29 obtained by Marimutu et a^10^ for India. Considering the rapid rate of rise of infections at that time, 1.29 does not look like a realistic estimate. Our calculations show that the reproduction number even rose to 10.08 later that week because of a sudden jump in cases at that time. This feature was not captured by this previous article on reproduction number. Ours is based on infectious period of 10 days but Marimutu et al. have not specified this period.

Despite such limitations of the study, we may not have faltered in our results for the reason already stated for each limitation.

## Conclusion

Testing rates seem to have an impact on the doubling time and the reproduction number - thus in controlling the epidemic. We now have evidence to recommend that the testing must be continuously increased at a fast pace along with the other measures. If the present trend continues, the doubling time will become 22.8 days and period reproduction number 0.89 when the testing reaches 30 per 1000 population. The period reproduction number is expected to become 1.0 when the testing is 20.3 per 1000 population. This may have already reached by the end of August 2020.

## Data Availability

All data availble at https://theconversation.com/how-long-are-you-infectious-when-you-have-coronavirus-135295

